# STM-GNN: Space-Time-and-Memory Graph Neural Networks for Predicting Multi-Drug Resistance Risks in Dynamic Patient Networks

**DOI:** 10.1101/2025.05.27.25327491

**Authors:** Damien Geissbuhler, Alban Bornet, Catarina Marques, André Anjos, Sónia Pereira, Douglas Teodoro

**Affiliations:** Department of Radiology and Medical Informatics, University of Geneva, Geneva, Switzerland; ciTechCare - Center for Innovative Care and Health Technology, Polytechnic University of Leiria, Leiria, Portugal; Idiap Research Institute, Martigny, Switzerland

**Keywords:** Hospital acquired infection, Temporal graph neural network

## Abstract

Hospital-acquired infections (HAIs), particularly those caused by multidrug-resistant (MDR) bacteria, pose significant risks to vulnerable patients. Accurate predictive models are important for assessing infection dynamics and informing infection prediction and control (IPC) programmes. Graph-based methods, including graph neural networks (GNNs), offer a powerful approach to model complex relationships between patients and environments but often struggle with data sparsity, irregularity, and heterogeneity. We propose the space-time-and-memory (STM)-GNN, a temporal GNN enhanced with recurrent connectivity designed to capture spatiotemporal infection dynamics. STM-GNN effectively integrates sparse, heterogeneous data combining network information from patient-environment interactions and internal memory from historical colonization and contact patterns. Using a unique IPC dataset containing clinical and environmental colonization information collected from a long-term healthcare unit, we show that STM-GNN effectively addresses the challenges of limited and irregular data in an MDR prediction task. Our model reaches 0.84 AUROC, and achieves the most balanced performance overall compared to classic machine learning algorithms, as well as temporal GNN approaches.

## 1 Introduction

Hospital-acquired infections (HAI), or nosocomial infections, are preventable adverse events in healthcare, yet they significantly increase healthcare mortality and costs [1]. High patient densities and extensive antibiotic use promote antimicrobial-resistant (AMR) bacteria, a major cause of HAI-related deaths [2, 3] and can persist in hospital environments for years [4]. In the absence of effective treatments, infection prevention and control (IPC) measures—identifying high-risk patients, isolating carriers, optimizing antibiotic use, and maintaining hygiene—are essential [5]. Reliable predictive models are crucial for evaluating IPC strategies and assessing individual infection risks, but prediction is challenging due to the diversity of HAIs [2] and their transmission routes, including direct contact, airborne spread, and asymptomatic carriers [6]. Hence, effective HAI modeling requires integrating patient history, inter-patient contact, and environmental factors.

Graph-based methods model HAI propagation by representing patients, healthcare workers, and environments as nodes, with interactions as edges that evolve as patients move through the hospital. Stochastic transmission models, assigning states to patients (e.g., susceptible, infected) and calculating transition rates, can be applied to IPC tasks like minimizing transmission [7], identifying infection sources [8], and detecting asymptomatic spreaders [6]. While these models provide an accurate framework for HAI dynamics, they rely solely on propagation pattern approximations and graph topology, which limits their generalizability across contexts [9].

Machine learning (ML) methods, supported by a growing corpus of digitized medical data, offer generalizable models for HAI prediction learning from electronic health records (EHRs) [10]. ML algorithms have been successfully applied to predict hospital-onset COVID infections, while remaining interpretable [11]. Graph neural networks (GNNs) are highly effective in epidemic modeling by aggregating network information and node features [9]. GNNs can leverage EHR-similarity graphs [12] or hospital interaction networks [13] to predict patient-level HAI risks.

Temporal graphs enhance GNNs by capturing the dynamic nature of hospital interactions and improving their predictive power with temporally dependent features [14, 15]. Strategies for temporal graphs include using LSTM modules for representing time-series in nodes [16] or adapting the message-passing algorithm dynamically [17]. Continuous-time dynamic graphs can combine GNNs with memory modules, updating node embeddings from aggregated graph events [18]. Alternatively, GNNs can be trained using static hyper-graphs derived from temporal graphs via snapshot concatenation [19] or by constructing causal graphs [20]. Still, challenges remain in applying ML algorithms and GNNs to temporal patient data, including data irregularity, heterogeneity, and sparsity, while also ensuring interpretable results [21].

In this work, we propose a deep learning approach, space-time-and-memory (STM)-GNN for MDR prediction using temporal graphs. STM-GNN integrates clinical and environmental colonization data to assess patient-level risks of MDR bacterial infections. The model implements a feedback loop between GNN and LSTM modules to capture both temporal and spatial factors through contact histories and measurements from connected samples. To address data sparsity and irregularity, node embeddings are augmented with temporal features before GNN processing, while node memory states incorporate spatial and network information prior to LSTM processing. The core idea is to contextualize model predictions by replaying patient trajectories within the evolving contact network. We evaluate STM-GNN against comparable ML and deep learning architectures using a proprietary dataset of bacterial smears collected in a long-term healthcare unit (LTHU). The data offer a realistic setting to assess MDR prediction performance. We show that:

- STM-GNN achieves a more balanced performance in predicting MDR infections compared to existing architectures in the low and imbalanced data regime.
- Ablation studies confirm that the feedback loop between GNN and LSTM modules is required to improve model predictions.

## 2 Methods

### 2.1 IPC Dataset

The proprietary dataset used in this study includes antibiotic sensitivity tests conducted on bacterial swab samples collected over six months in a long-term healthcare unit (LTHU). Ethical approval for this study was obtained from the Ethics Committee of the Polytechnic of Leiria (CE/IPLEIRIA/40/2020).

Swab samples were collected biweekly and for 6 months from 17 patients (mouth, hands, anus), 14 beds (bedside table, bed railing, handbell), and 9 rooms (bathroom door handle). Samples were also collected under various study conditions (admission, transfer, discharge, room cleaning). The dataset distinguishes between clinical samples, collected directly from patients, and environmental samples, taken from beds and rooms. Samples were screened for bacterial growth and processed into cultures, which were tested for MDR. Each microbiological test result from swab samples is linked to a unique identifier for the corresponding patient, bed, or room, and associated with the date the sample was collected. Figure 1 shows the timeline of all samples with their colonization status.

**Fig. 1.**
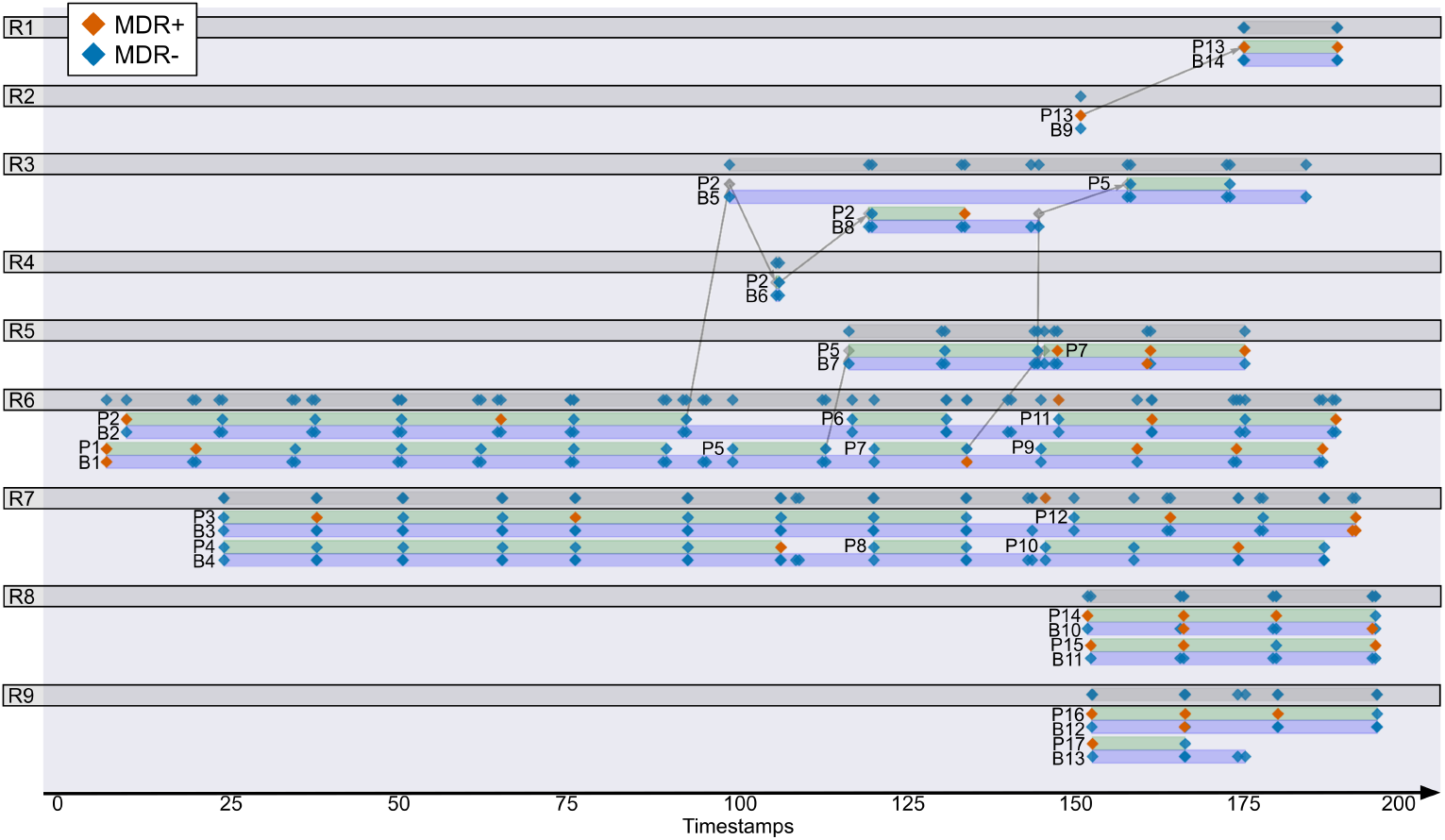
Sampling history across rooms, beds, and patients. Each diamond represents a sampling event, colored red any swab samples detected an MDR strain, blue other-wise. Gray lines represent patient transfers within the LTHU. Patient, bed, and room numbers were renumbered based on their order of appearance.

### 2.2 Temporal Graph Construction

To model the dynamics of MDR colonizations, we created a temporal graph by mapping our dataset to a sequence of static heterogeneous graph snapshots. We built the static graphs by grouping samples sharing the same sampling date, study condition, room, and cleaning occurrence and arranged them chronologically according to the sampling event date. Graphs were assembled by identifying patient, bed, and room nodes present within each sample groups. We defined an heterogeneous graph by considering two node types: clinical (patients) and environmental (beds, rooms). To capture the spatial and contextual relationships of the environment, edges connected any pair of patients present simultaneously in the same room. Moreover, we added edges linking each patient node to its assigned bed and room.

Each node was assigned the following static and time-dependent features. Static features included patient vitals (blood pressure, heart rate, temperature, oxygen saturation, blood sugar level), patient hospital trajectory (admission, continuity, post-discharge sampling, room transfers), demographic details (age, weight, height, medical history, current condition), as well as room occupancy and area. Time-dependent variables included patient’s length of stay (in days), the interval between consecutive samples from the same location, and the number of healthcare workers (HCWs) involved in cleaning. Finally, we derived the colonization pressure [22, 23] as the proportion of carriers among admitted patients over the past 30 days, and defined a boolean indicator for whether any samples from the same location in the last 30 days tested positive for MDR bacteria.

Finally, nodes were assigned MDR target labels. Patient nodes were labeled positive if at least one swab sample from the hand, mouth, or anus yielded a culture positive for MDR. Bed nodes were labeled similarly, based on the results of the samples from bed railing, hand-bell, and bedside table. Room nodes were labeled positive if the bathroom door handle sample tested positive for MDR. Using this method, the dataset was mapped to 132 static graph snapshots, including 372 node observations and 668 edges. Table 1 shows node statistics for each MDR label.

**Table 1.**
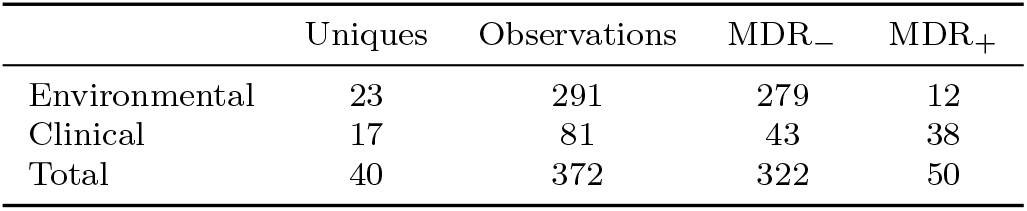
Node counts and MDR distribution.

|  | Uniques | Observations | MDR<sub>-</sub> | MDR<sub>+</sub> |
| --- | --- | --- | --- | --- |
| Environmental | 23 | 291 | 279 | 12 |
| Clinical | 17 | 81 | 43 | 38 |
| Total | 40 | 372 | 322 | 50 |

### 2.3 STM-GNN Model

We propose the space-time-and-memory (STM)-GNN architecture, designed under the assumption that MDR bacterial colonization is strongly influenced by the contact history between patients and their environment. The model processes a temporal graph as a sequence of snapshots and produces node embeddings for each snapshot. To handle heterogeneous graphs and accommodate long memory sequences, we modified the Temporal Graph Networks (TGN) model [18].

At each time step, a linear layer embeds the nodes features of the snapshot graph being processed. Then, the STM-GNN model processes node embeddings using two core modules, which jointly act to capture the temporal evolution of the nodes given their connectivity. Figure 2 shows the flow of operations performed by the model on a single graph snapshot.

**Fig. 2.**
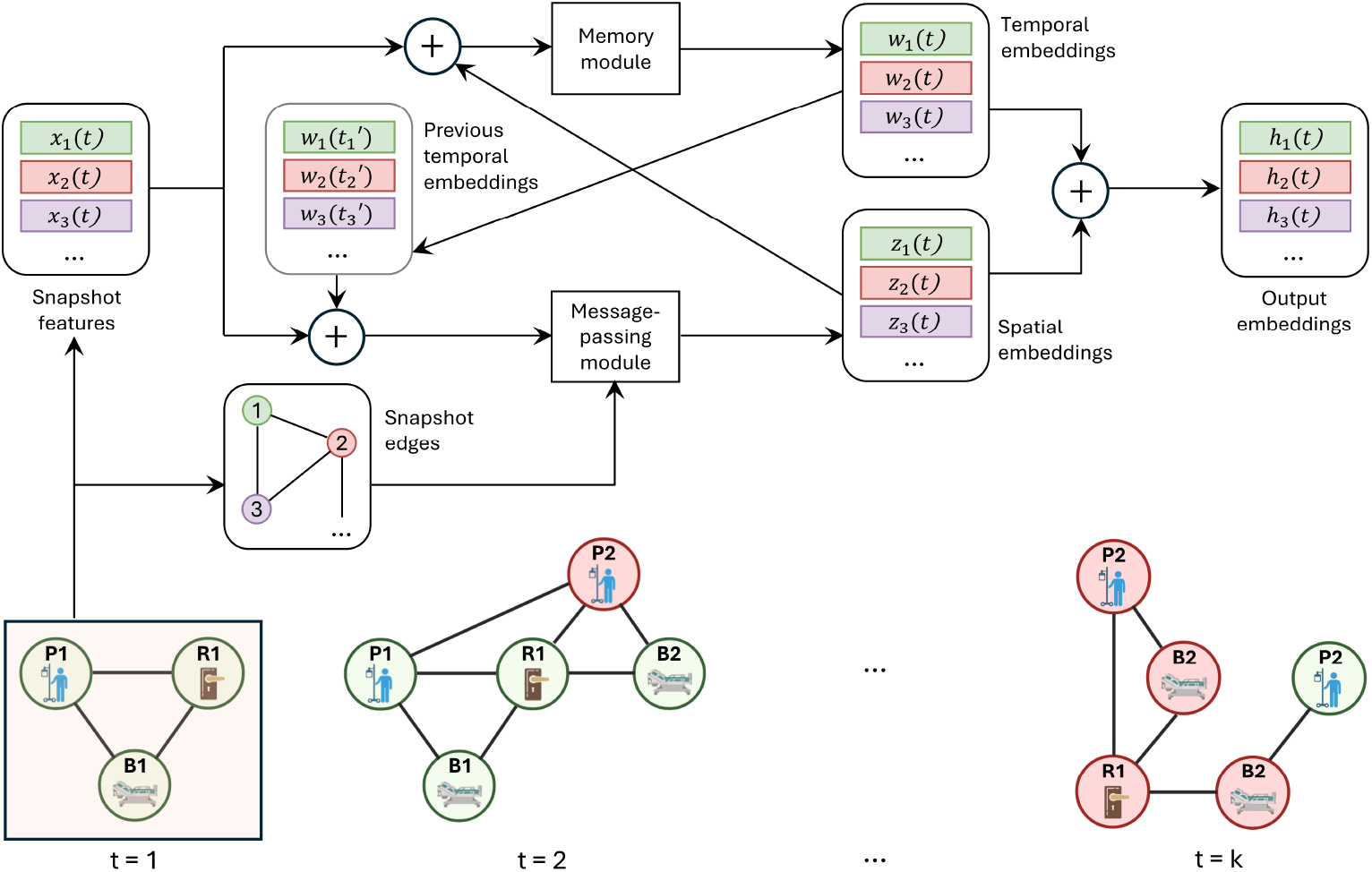
Workflow of the STM-GNN model applied to a single snapshot graph within a sequence. Temporal embeddings from previous time steps are retrieved from memory and combined with current graph node features. Message passing generates updated node embeddings, which are added to the original node features and stored in memory. The memory module produces time-series embeddings, which are summed with spatial embeddings to derive spatiotemporal representations.

#### Memory Module

First, the memory module gathers the history of each node by concatenating the embedding vectors of incoming nodes into a memory state *M*_*i*_(*t*_*k*_), from which temporal node embedding are derived via an aggregation function *agg*:

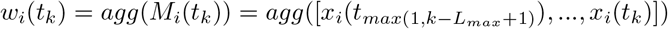

*M*_*i*_(*t*_*k*_) is a matrix of shape *L*_*k*_ *×d*, where *d* is the embedding dimension and *L*_*k*_ *≤ L*_*max*_ the number of time steps in memory, with *L*_*max*_ the memory capacity. We used an attention layer as the aggregation function.

#### Message Passing Module

Given a graph snapshot *G*(*t*), spatial node embedding are computed from node features and edge information using the message passing algorithm, with the general form:

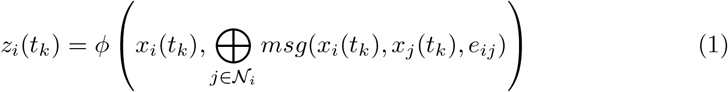

where *ϕ* and *msg* are learnable functions, and ⊕ EB is any permutation invariant aggregation operator. To implement *ϕ, msg*, and ⊕, we used the graph attentional network (GAT) framework [24].

#### Recurrent Augmentations

To enhance the integration of temporal and spatial contexts into node embeddings, the STM-GNN model implements recurrent augmentations. For each snapshot graph, node features are enriched by adding previous temporal embeddings before they are processed by the message-passing module. Similarly, spatial embeddings are added to node features before storing them in memory and processing them with the temporal aggregation module.

### 2.4 Experimental Setting

All models were trained using a nested cross-validation scheme [25], where the temporal graph was split into 7-day intervals. In each fold, the last sequence served as the test set and the remaining sequences were used for training and validation. Binary cross-entropy was minimized using a weighted loss function to address class imbalance across node types. The pooled size of each fold’s test set is 108 nodes, which ensures 0.99 power, 0.05 significance level, with an effect size of 0.2 and a baseline proportion of 0.134. Data preprocessing included robust scaling for numerical features, one-hot encoding for categorical features, and imputation based on sample statistics. We compared STM-GNN against several baselines: (i) classic ML models trained independently on node features for each node type; (ii) temporal-only models using LSTM with attention (ATTN) on node time-series; (iii) spatial-only models leveraging the graph structure of individual snapshots with methods like GAT and GraphSAGE (GSAGE); and (iv) a spatiotemporal graph model (TGN) that incorporates both temporal and structural information but ignores node heterogeneity. We used Optuna (100 trials) to tune the hyper-parameters of all models, with balanced accuracy within the nested cross-validation framework as the objective metric.

## 3. Results

Table 2 summarizes the performance metrics obtained with all models on the test dataset, with 95% confidence intervals obtained through bootstrapping with 1,000 resamples. STM-GNN achieves the best set of performance metrics overall. Moreover, ablation study results show that removing spatial and/or temporal augmentations reduces most metrics. Figure 3 shows ROC and PR curves, as well as STM-GNN’s performance stratified by sample type.

**Table 2.** MDR infection prediction test performance obtained with STM-GNN and comparative baseline models, as well as ablation studies (T-: no temporal augmentation; S-: no spatial augmentation; ST-: no augmentation).

| Model | Accuracy | Sensitivity | Specificity | F1-score | AUROC | AP |
| --- | --- | --- | --- | --- | --- | --- |
| LR | 35.8 ± 9.0 | 73.9 ± 18.7 | 25.3 ± 9.3 | 33.3 ± 12.0 | 46.5 ± 14.3 | 24.6 ± 13.1 |
| XGB | 75.5 ± 8.5 | 34.8 ± 19.8 | 86.7 ± 7.6 | 38.1 ± 18.8 | 68.9 ± 13.6 | 45.2 ± 18.0 |
| SVM | 74.5 ± 8.0 | 26.1 ± 18.2 | 88.0 ± 6.7 | 30.8 ± 19.3 | 62.3 ± 12.4 | 32.7 ± 15.6 |
| LSTM | 47.2 ± 9.4 | 43.5 ± 19.9 | 48.2 ± 10.6 | 26.3 ± 13.5 | 50.2 ± 13.4 | 30.1 ± 16.1 |
| ATTN | 85.8 ± 6.6 | 56.5 ± 20.1 | 94.0 ± 5.2 | 63.4 ± 17.8 | 77.3 ± 9.1 | 45.3 ± 19.0 |
| GSAGE | 83.0 ± 7.1 | 47.8 ± 20.7 | 92.8 ± 5.1 | 55.0 ± 18.3 | 82.1 ± 9.5 | 56.8 ± 19.1 |
| GAT | 72.6 ± 8.5 | 30.4 ± 18.8 | 84.3 ± 7.6 | 32.6 ± 18.5 | 79.9 ± 10.7 | 62.3 ± 18.2 |
| TGN | 72.6 ± 8.5 | 0.0 ± 0.0 | 92.8 ± 5.5 | 0.0 ± 0.0 | 64.7 ± 11.8 | 28.9 ± 13.1 |
| STM | 86.8 ± 6.6 | 43.5 ± 20.8 | 98.8 ± 2.2 | 58.8 ± 20.6 | 84.4 ± 9.8 | 71.3 ± 15.6 |
| STM-T- | 83.0 ± 7.1 | 43.5 ± 20.4 | 94.0 ± 5.3 | 52.6 ± 19.6 | 79.0 ± 11.0 | 64.0 ± 16.8 |
| STM-S- | 49.1 ± 9.4 | 43.5 ± 19.6 | 50.6 ± 10.6 | 27.0 ± 13.6 | 60.5 ± 14.1 | 42.5 ± 18.4 |
| STM-ST- | 85.8 ± 6.6 | 47.8 ± 20.6 | 96.4 ± 3.9 | 59.5 ± 19.0 | 80.2 ± 11.8 | 65.6 ± 18.0 |

**Fig. 3.**
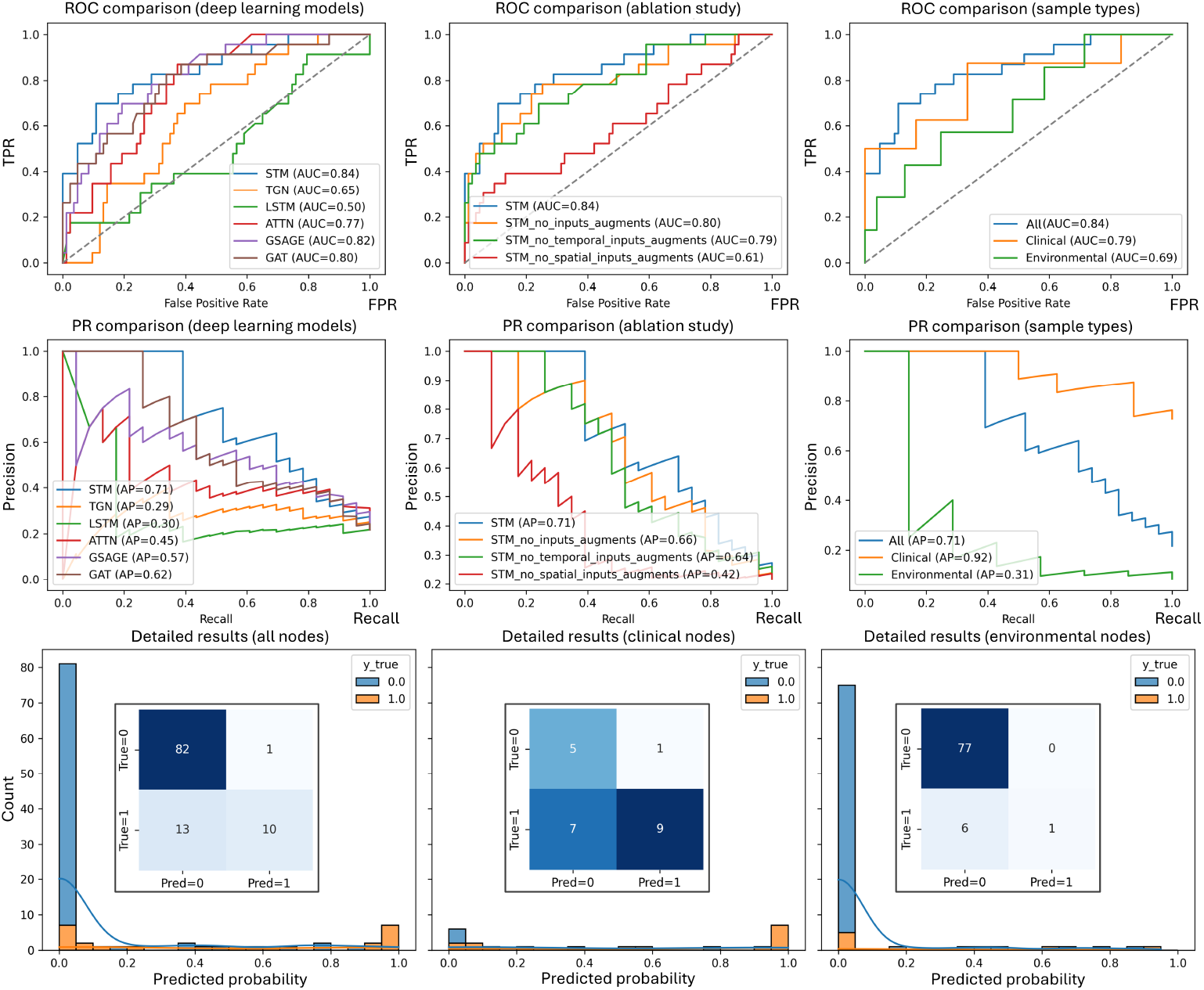
ROC (top) and PR (middle) curve comparisons for deep learning models (left), for different ablations (center), and stratified by sample type (center). MDR prediction confusion matrix and probability histogram for different sample types (bottom).

## 4. Discussion

We propose the STM-GNN model, a novel approach for predicting MDR infections by integrating spatial and temporal dependencies through recurrent augmentations. Our findings show that STM-GNN provides a more balanced and robust performance compared with classical ML methods and existing deep or graph-based approaches (Table 2). The ablation study suggests that the integration of recurrent augmentations between temporal memory and spatial embeddings contributes to this improvement (Figure 3, center). Interestingly, STM-GNN without spatial augmentation obtains a similar (and similarly low) pattern of performance as the LSTM model, which confirms that recurrent connectivity is necessary for network interactions and spatial relationships to improve prediction accuracy beyond simple temporal memory.

Stratifying results by sample type (Figure 3, bottom) shows that the STM-GNN model more accurately predicts MDR status in clinical samples, where positives are relatively common, than in environmental samples, where positives are proportionally rare due to IPC measures such as room cleaning conducted during data collection. As a result, the model tends to “specialize” in identifying clinical positives and under-estimate the risk in environmental locations, translating into higher specificity but reduced sensitivity. This behavior is expected given the class imbalance (see Table 2, where most models exhibit low sensitivity, particularly the TGN model, which does not handle node heterogeneity), yet STM-GNN achieves the best trade-off despite the limited data. Addressing this imbalance by expanding the dataset or ensuring a more even distribution across clinical and environmental samples could improve model performance.

Future work could explore alternative graph constructions, such as sample-based graphs rather than patient-, bed-, and room-based graphs, where connectivity is defined by biologically relevant transmission pathways (e.g., hand-to-bed railing interactions but not mouth-to-door handle contacts). Moreover, healthcare worker links could enhance the temporal graphs with inter-rooms connection. Finally, STM-GNN could be validated through synthetic data generated using known infection models (e.g. SIR) to control feature dependence and test the capacity of the model to reproduce different transmission routes for different levels of data sparsity and irregularity. The model could also be evaluated with publicly available patient interaction graphs, such as MIMIC, where random time shifts make the derived graphs effectively synthetic.

## Data Availability

The data that support the findings of this study are not publicly available due to their proprietary nature and patient privacy restrictions.

